# Prolonged SARS-CoV-2 infection in patients with lymphoid malignancies

**DOI:** 10.1101/2021.08.25.21262417

**Authors:** Christina Y. Lee, Monika K. Shah, David Hoyos, Alexander Solovyov, Melanie Douglas, Ying Taur, Peter G. Maslak, N. Esther Babady, Benjamin Greenbaum, Mini Kamboj, Santosha A. Vardhana

**Affiliations:** Lymphoma Service, Division of Hematologic Malignancies, Department of Medicine, Memorial Sloan Kettering Cancer Center, New York, NY, USA; Department of Medicine, Weill Cornell Medical College, Cornell University, New York, NY, USA; Infectious Diseases Service, Department of Medicine, Memorial Sloan Kettering Cancer Center, New York, NY, USA; Computational Oncology, Department of Epidemiology and Biostatistics, Memorial Sloan Kettering Cancer Center, New York, NY, USA; Department of Medicine, Memorial Sloan Kettering Cancer Center; Department of Laboratory Medicine, Memorial Sloan Kettering Cancer Center; Clinical Microbiology Service, Department of Laboratory Medicine, Memorial Sloan Kettering Cancer Center, New York, NY, USA; Physiology, Biophysics & Systems Biology, Weill Cornell Medicine, Weill Cornell Medical College, New York, NY, USA; Infection Control, Division of Quality and Safety, Memorial Sloan Kettering Cancer Center, New York, NY, USA; Human Oncology and Pathogenesis Program, Memorial Sloan Kettering Cancer Center, New York, NY, USA; Parker Institute for Cancer Immunotherapy, San Francisco, CA, USA

## Abstract

Coronavirus disease 2019 (COVID-19) infection results in high mortality rates in patients with hematologic malignancies. Persistent and/or recurrent COVID-19 has not yet been demonstrated in this population. We identified patients with B-cell lymphomas as having a particularly high risk for persistent SARS-CoV-2 positivity. Subsequent analysis of patients with lymphoid malignancies and COVID-19 identified discrete risk factors for severity of primary infection as compared to disease chronicity. Active therapy and diminished T-cell counts were key drivers of acute mortality in lymphoma patients with COVID-19 infection. Conversely, B-cell depleting therapy was the primary driver of re-hospitalization for COVID-19. In patients with persistent SARS-CoV-2 positivity, we observed high levels of viral entropy consistent with intrahost viral evolution, particularly in patients with impaired CD8+ T-cell immunity. These results suggest that persistent COVID-19 infection is likely to remain a risk in patients with impaired adaptive immunity and that additional therapeutic strategies are needed to enable viral clearance in this high-risk population.

**Statement of Significance:** We establish persistent symptomatic COVID-19 infection as a novel clinical syndrome in patients with lymphoid malignancies and identify B-cell depletion as the key immunologic driver of persistent infection. Furthermore, we demonstrate ongoing intrahost viral evolution in patients with persistent COVID-19 infection, particularly in patients with impaired CD8+ T-cell immunity.

## INTRODUCTION

Severe acute respiratory syndrome coronavirus 2 (SARS-CoV-2) infection can lead to chronic manifestations in healthy and immunocompromised individuals with different clinical presentations and unique risks. In otherwise healthy people hospitalized with coronavirus disease 2019 (COVID-19), up to one-third continue to experience physical and cognitive symptoms, a syndrome commonly referred to as long COVID.^1^ In severely immunocompromised hosts, protracted respiratory symptoms with frequent readmissions can occur. The underlying pathogenic mechanisms for the two entities are not well understood. However, for severely immunocompromised patients, isolated cases of persistent COVID-19 clinical manifestations with co-incident persistent respiratory SARS-CoV-2 RNA detection and the absence of seroconversion have been reported.^2,3^

Certain immunocompromising conditions, such as hematologic malignancy, are associated with a higher risk for severe COVID-19 and poor outcomes after acute infection.^4–6^ The unusual and protracted disease course marked by intermittent flares is less defined but a frequently encountered COVID-19 complication in this patient population. Persons with lymphoid malignancy, especially those with diminished T-cell responses and after B-cell depleting therapies, are particularly susceptible.^7^ A few case reports describe the clinical and viral genomic evolution in chronically infected patients with lymphoid malignancies and highlight the immunologic correlation with impaired CD4 and CD8 virus-specific T-cell responses.^3,8^

In addition to the substantial direct morbidity from COVID-19 and interruption in cancer care, chronic persistent SARS-CoV-2 infection poses the additional threat of extended transmissibility and selection pressure that could generate immune evading mutations.^8,9^ Intra-host evolution in chronic SARS-CoV-2 infection has led to the detection of mutations found in variants of concern (VOC), raising intense speculation that immunocompromised patients may serve as reservoirs for the emergence of critical mutations in the receptor binding domain (RBD) region of the virus.^8^

Therefore, a better understanding of chronic infection in severely immunocompromised patients is significant from both a therapeutic and public health perspective. In the present study, we identify predictors of persistent COVID-19 among patients with hematologic malignancies and characterize the clinical course and viral genomic evolution in patients with lymphoid malignancy, including those receiving B-cell depleting therapies.

## RESULTS

### Persistent SARS-CoV-2 reverse-transcriptase–polymerase-chain reaction (RT-PCR) positivity in patients with hematologic malignancy

From March 10, 2020 until February 28, 2021, 382 patients with hematologic malignancy were diagnosed with COVID-19, including 214 patients with lymphoid malignancies (**Figure 1**). Persistent PCR positivity, defined as SARS-CoV-2 RNA detection ≥30 days after initial positivity, was detected in 51 (13.9%) of 368 patients alive at 30 days from COVID-19 diagnosis. The demographic and clinical features of the 368 patients as well as univariate and multivariate predictors of persistent PCR positivity are shown in **Supplementary Table 1 and Table 1**, respectively. Receipt of systemic parenteral chemotherapy within 30 days, treatment with anti-CD20 antibodies within 1 year, and cellular therapy including hematopoietic stem cell transplantation (HSCT) within 1 year were independent predictors of prolonged SARS-CoV-2 RNA detection. Age and underlying lymphoid malignancy did not independently influence prolonged viral detection among patients with hematologic cancers.

**Table 1:**
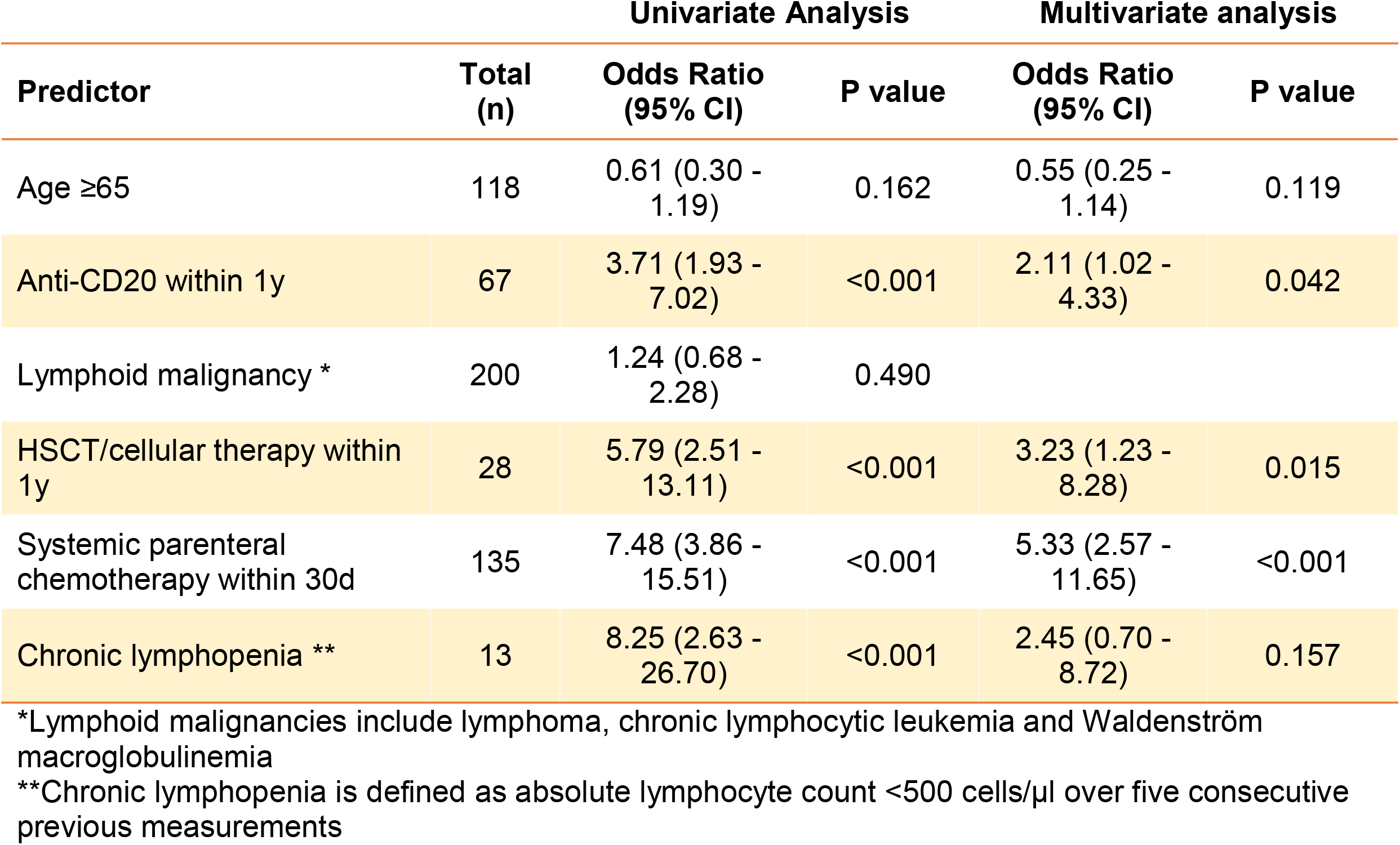
Predictors of SARS-CoV-2 persistence in patients with hematologic cancers.

| Predictor | Total (n) | Univariate Analysis |  | Multivariate analysis |  |
| --- | --- | --- | --- | --- | --- |
|  |  | Odds Ratio (95% CI) | P value | Odds Ratio (95% CI) | P value |
| Age ≥65 | 118 | 0.61 (0.30 - 1.19) | 0.162 | 0.55 (0.25 - 1.14) | 0.119 |
| Anti-CD20 within 1y | 67 | 3.71 (1.93 - 7.02) | <0.001 | 2.11 (1.02 - 4.33) | 0.042 |
| Lymphoid malignancy * | 200 | 1.24 (0.68 - 2.28) | 0.490 |  |  |
| HSCT/cellular therapy within 1y | 28 | 5.79 (2.51 - 13.11) | <0.001 | 3.23 (1.23 - 8.28) | 0.015 |
| Systemic parenteral chemotherapy within 30d | 135 | 7.48 (3.86 - 15.51) | <0.001 | 5.33 (2.57 - 11.65) | <0.001 |
| Chronic lymphopenia ** | 13 | 8.25 (2.63 - 26.70) | <0.001 | 2.45 (0.70 - 8.72) | 0.157 |

\*Lymphoid malignancies include lymphoma, chronic lymphocytic leukemia and Waldenström macroglobulinemia
\*\*Chronic lymphopenia is defined as absolute lymphocyte count <500 cells/μl over five consecutive previous measurements

**Figure 1:**
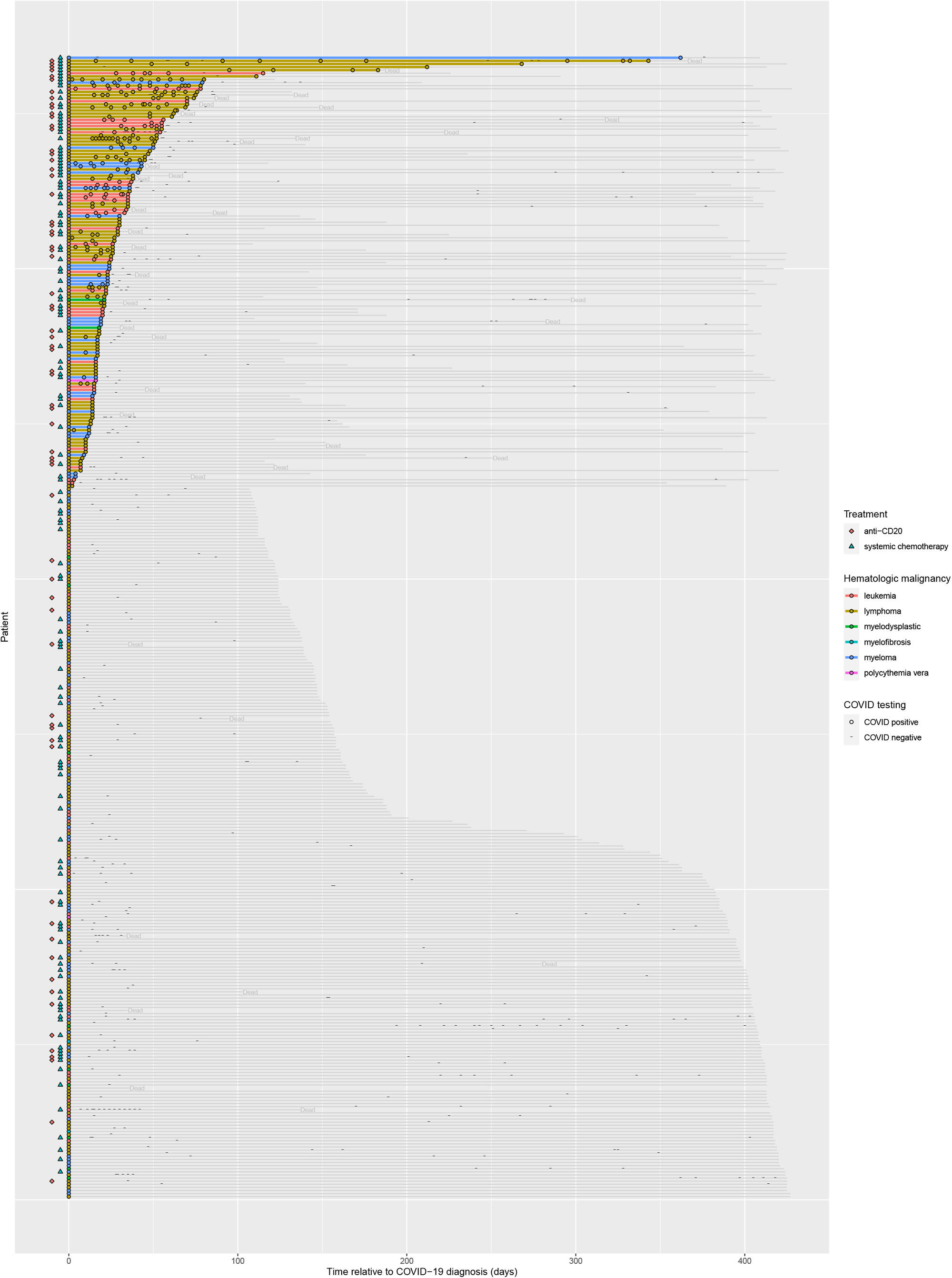
High rates of persistent SARS-CoV-2 PCR positivity in hematologic malignancy patients who received systemic chemotherapy or B-cell depleting agents. Swimmer’s plot depicting duration of known SARS-CoV-2 PCR positivity relative to first known positive PCR test. Grey lines indicate duration of follow-up. Colored lines represent times of persistent positivity (at least two positive tests). Treatments, hematologic cancer subtypes, deaths during follow-up, and individual PCR test results as indicated.

### SARS-CoV-2 infection in patients with lymphoid cancers

Given that the strongest predictors of viral persistence are standard of care therapies for patients with lymphoid malignancies, we subsequently identified 214 patients with lymphoid malignancy and COVID-19 infection between March 2020 and January 2021 (**Supplementary Table 2**). The majority of patients (86%) had non-Hodgkin lymphoma, of which the most common subtypes were chronic lymphocytic leukemia/small lymphocytic lymphoma (20%), diffuse large B-cell lymphoma (19%), and follicular lymphoma (15%). Approximately half of the patients had received recent systemic therapy, including anti-CD20 monoclonal antibodies (32%), Bruton tyrosine kinase (BTK) inhibitors (10%), autologous or allogeneic stem cell transplantation (7%), or CD19-directed chimeric antigen receptor (CAR) T-cell therapy (3%). Seventy-nine patients (37%) were undergoing active management at the time of COVID-19 diagnosis, while 82 patients (38%) were under surveillance and 53 patients (25%) were in remission.

Most patients (83%) met criteria for severe COVID-19, with 73 of 102 (72%) hospitalized patients requiring supplemental oxygen, 32 of 102 (31%) requiring intensive care unit (ICU)-level care, and 22 of 102 (22%) requiring mechanical ventilation. The management and outcomes of COVID-19 in inpatients are summarized in **Supplementary Tables 3 and 4**. Inpatients were treated with hydroxychloroquine (40%) in March and April 2020, convalescent plasma (44%), remdesivir (37%), dexamethasone (24%) and/or other systemic steroids (30%), intravenous immune globulin (9%), tocilizumab (7%), anakinra (1%), and N-acetylcysteine (17%) on a clinical trial (ClinicalTrials.gov Identifier: NCT04374461). The case fatality rate during the index hospitalization was 22%. Thirty-two patients (31%) experienced an extended hospitalization over 21 days, and 19 out of the remaining 77 patients who were discharged following index hospitalization were readmitted with recrudescent respiratory symptoms after initial improvement. Most of these patients had continuous COVID-19 PCR positivity, suggestive of a high rate of incomplete viral clearance in this population.

### Clinical and immunological hallmarks of severe acute SARS-CoV-2 infection in patients with lymphoma

Univariate analysis identified clinical and laboratory features associated with death from acute COVID-19 infection, as shown in **Figure 2a** and **Supplementary Figure 1a**. Key predictors of death included cardiovascular illness, relapsed/refractory disease, and active treatment, including anti-CD20 and CAR T-cell therapy in the past year. Multivariate analysis revealed cardiovascular disease, active treatment, and CAR T-cell therapy, but not anti-CD20 therapy, to be independently associated with mortality.

**Figure 2:**
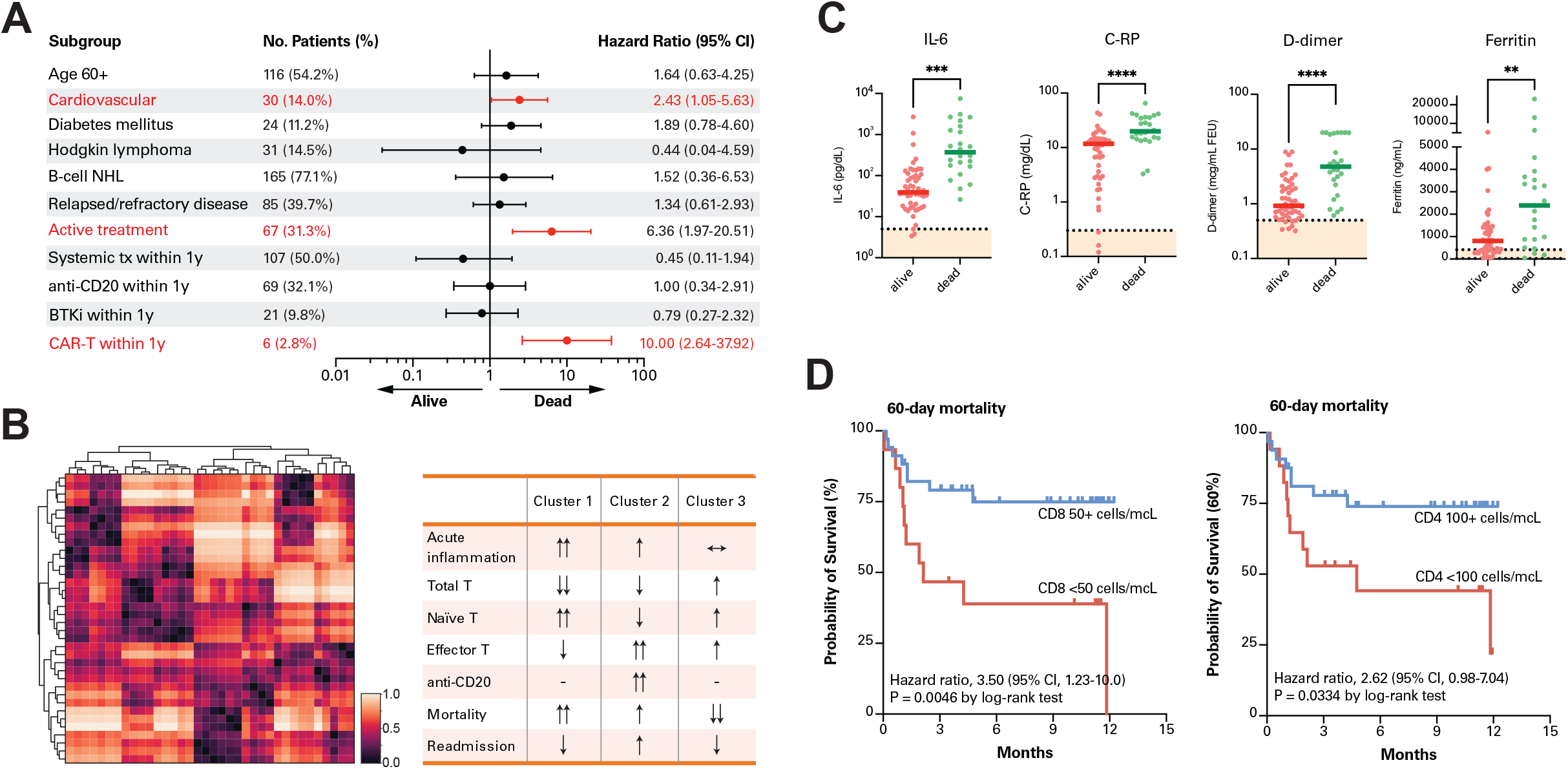
Predictors of mortality during acute SARS-CoV-2 infection in patients with lymphoma. a) Multivariate analysis of clinical risk factors predicting mortality attributable to COVID-19 infection in patients with lymphoma. Statistically significant risk factors highlighted in red. b) Left, two-dimensional Uniform Manifold Approximation and Projection (UMAP) of laboratory data for 36 patients with lymphoma in whom at least 10% of all laboratory analyses were conducted. Missing data is imputed. Right, description of three patient clusters generated based on Earth Mover’s Distance (EMD) of each patient relative to each other in UMAP space. c) Laboratory markers in patients who were alive or dead following acute COVID-19 infection. d) Kaplan-Meier curve displaying probability of survival within 60 days of acute COVID-19 infection in patients with decreased CD8+ or CD4+ T-cell counts. **p<0.01. ***p<0.001. ****p<0.0001.

Additionally, unbiased clustering of immunologic laboratory data obtained from patients during acute infection generated three semi-global clusters of patients with distinct immunologic phenotypes (**Figure 2b** and **Supplementary Figure 2**). Cluster 1 was defined by the most significantly reduced levels of T-cells as well as the lowest ratio of effector or activated T-cells to naïve T-cells, but the highest levels of acute inflammatory markers, consistent with a blunted T-cell response. Cluster 2 was defined by intermediate T-cell counts, a relatively high incidence of anti-CD20 therapy, and a relatively high level of T-cell activation as well as moderate levels of acute inflammatory markers, consistent with a partially compensated immune response in the presence of B-cell depletion. Finally, cluster 3 was defined by minimal acute inflammatory markers, normal T-cell counts, and higher levels of T-cell activation consistent with a sufficient adaptive immune response. Correlation of immune clusters with COVID-specific clinical outcomes demonstrated the highest mortality in cluster 1 (5/11, 45%), the lowest mortality rate in cluster 3 (3/17, 18%), and intermediate mortality (3/8, 38%), but a high rate of readmission to the hospital for persistent symptomatic disease (4/8, 50%) in cluster 2.

We therefore asked whether specific inflammatory markers were associated with clinical outcomes in COVID-19-infected lymphoma patients. Hospitalized patients with lymphoma demonstrated similar immunologic profiles to those reported in the broader population, with elevated levels of all inflammatory markers and lymphopenia in approximately half (55%) of patients. Notably, however, severe lymphopenia (absolute lymphocyte count [ALC] less than 500 cells/µL) was present in 27% of hospitalized patients, and flow cytometry analysis revealed undetectable B-cell counts in 57% (25/44) of patients. Laboratory parameters that correlated with mortality included interleukin-6 (IL-6), C-reactive protein (C-RP), ferritin, D-dimer, and lactate dehydrogenase (LDH) (**Figure 2c** and **Supplementary Figure 1b**). Conversely, median levels of CD19^+^ B-cells, CD4^+^ T-cells, and CD8^+^ T-cells did not correlate with mortality from acute infection. (**Supplementary Figure 1b**). However, patients with either severe CD8^+^ lymphopenia (<50 cells/µL) or CD4^+^ lymphopenia (<100 cells/µL) had markedly poor outcomes (**Figure 2d**).

### Clinical and immunological hallmarks of chronic SARS-CoV-2 infection in patients with lymphoma

Given the high rate of persistent PCR positivity, protracted hospital stay and re-hospitalization in this population, we next asked whether distinct clinical or immunologic factors were associated with persistent COVID-19 infection. To uncouple factors associated with prolonged disease from factors associated with severe initial disease, we focused on the outcomes of patients whose COVID-19 was sufficiently severe to require hospitalization (n=102) to determine whether patients who died during their primary hospitalization (n=26), patients who were discharged and not re-admitted to the hospital (n=57), and patients who were re-admitted to the hospital for COVID-19-related symptomatology (n=19) exhibited distinct clinical and laboratory manifestations (**Supplementary Figure 3a**). The clinical description of these 19 cases is presented in **Table 2**. Notably, 13 of 19 patients were on active treatment and 15 had received anti-CD20 therapy in the preceding year. Five patients were allogenic HSCT or CAR-T recipients and were in remission at the time of COVID-19 diagnosis. The median number of readmissions was 2 (range 2–9), and the median time from diagnosis to first readmission was 36.5 days (interquartile range 38 days). The initial presentation at the time of diagnosis was characterized as asymptomatic in 3 patients, mild in 13, moderate in 2, and severe in 1. In between hospitalizations, all nineteen cases had recurrent, persistent or progressive respiratory symptoms accompanied by new fever in 53% of cases. Computed tomography (CT) demonstrated progressive ground glass pulmonary infiltrates in all cases without a clear alternate etiology (**Supplementary Figure 4**). The median duration of PCR positivity was 56 days (range 1–344 days). During a median follow up of 277 days, thirteen (68%) individuals with persistent COVID-19 manifestations required discontinuation, interruption, or modification of lymphoma-directed therapy (**Table 2**). Five patients died during the follow-up period; four deaths were attributed to progressive respiratory decline due to symptomatic COVID-19 and one death was attributed to both progressive symptomatic COVID-19 and concurrent pulmonary coccidiomycosis.

**Table 2:**
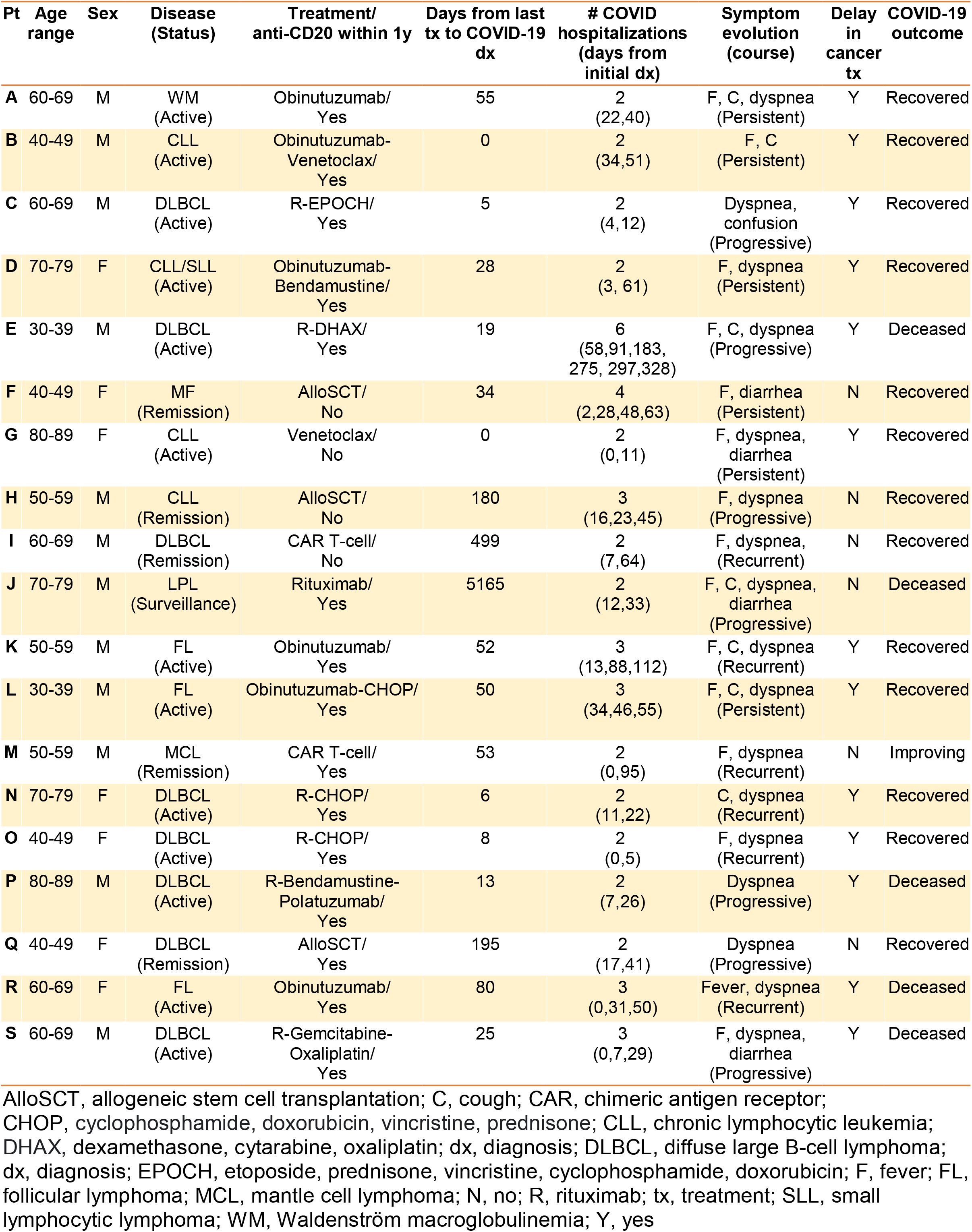
Baseline characteristics, clinical course, and outcomes of patients with chronic symptomatic SARS-CoV-2 infection requiring multiple hospital admissions.

**Figure 3:**
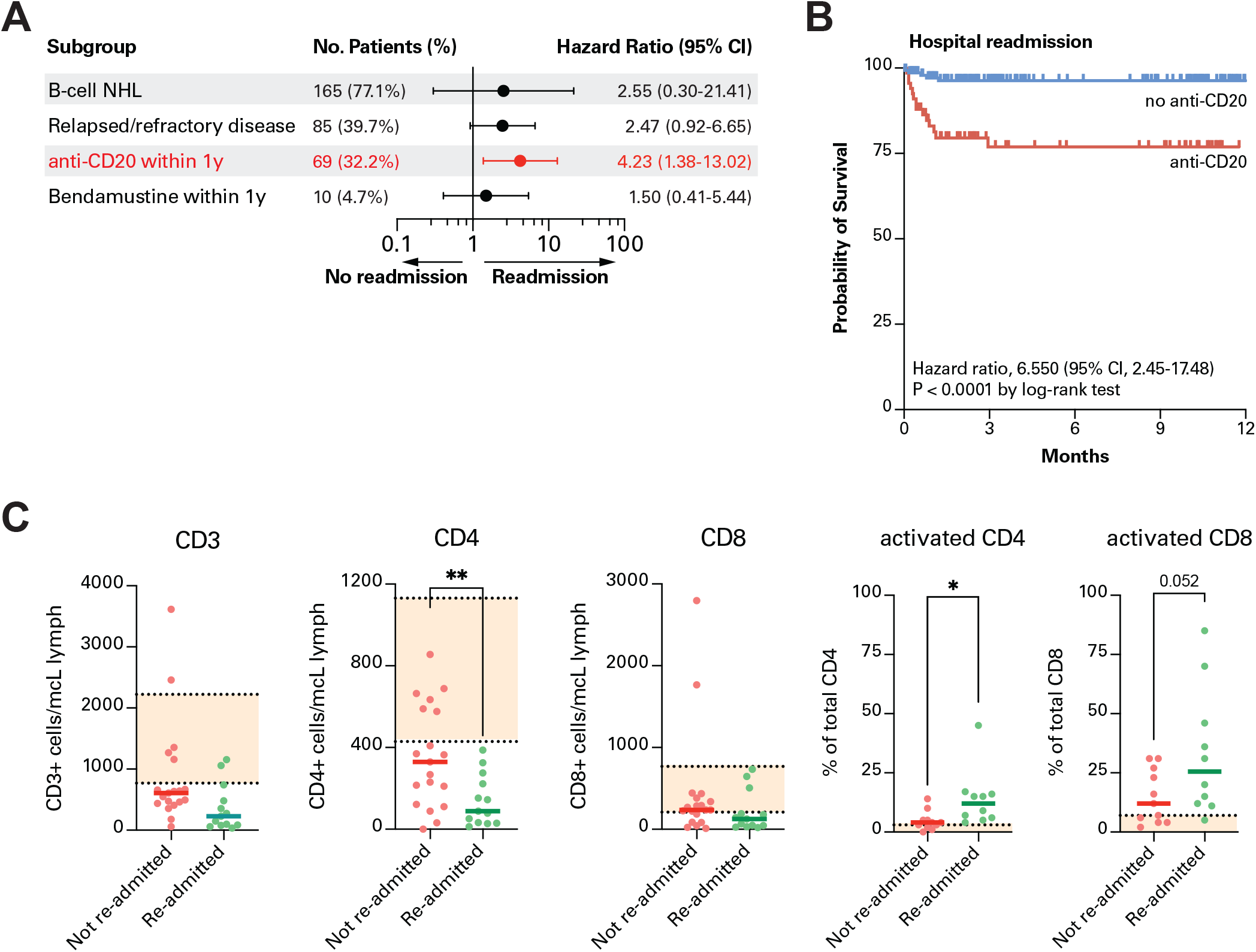
Adaptive immune dysfunction in lymphoma patients with recurrent COVID-19 infection. a) Multivariate analysis of clinical risk factors predicting re-hospitalization attributable to COVID-19 infection in patients with lymphoma and prior hospitalization attributable to COVID-19. Statistically significant risk factors highlighted in red. b) Kaplan-Meier curve displaying probability of re-hospitalization for COVID-19 in patients with lymphoma based on receipt of anti-CD20 therapy in the year prior to first known documented SARS-CoV-2 PCR positivity. c) Laboratory markers in patients with lymphoma who were either re-admitted or not re-admitted to the hospital for symptomatic COVID-19 infection following discharge from the hospital for acute COVID-19 infection. *p<0.05. **p<0.01.

**Figure 4:**
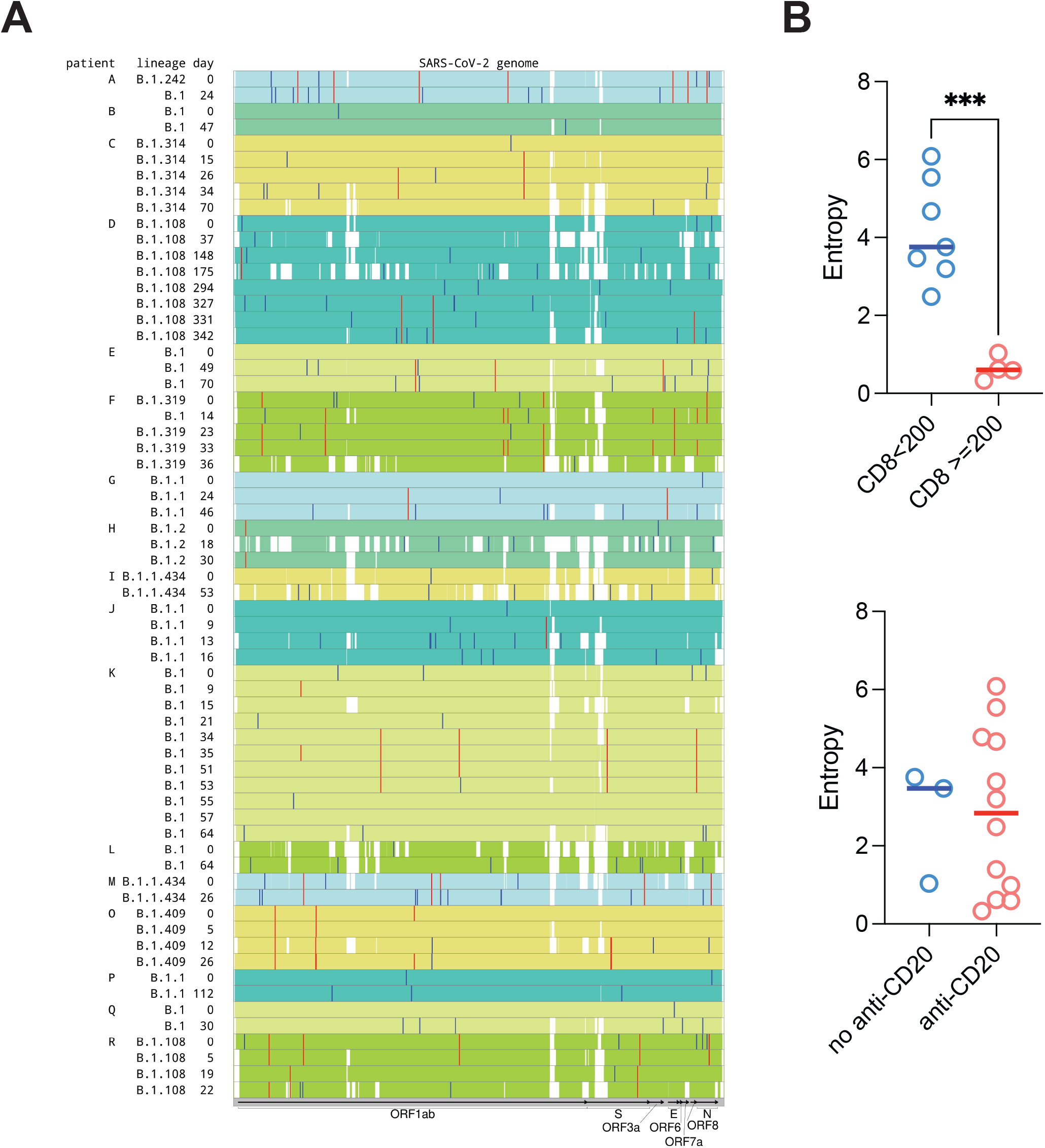
Increased SARS-CoV-2 viral genome entropy in patients with adaptive immune dysfunction. a) Minor variants in SARS-CoV-2 sequences from 17 patients with longitudinal data. Samples from the same patient are grouped together and highlighted with the same background color. White gaps indicate regions with low sequencing coverage. Minor variants. Location of each variant is indicated by a vertical line. Red lines represent the recurrent variants, which are seen in at least two samples from the same patient. Blue lines represent the variants unique to a particular sample. b) Viral entropy of the SARS-CoV-2 genome in patients with normal or decreased CD8^+^ T-cell counts (above), or patients who had or had not received anti-CD20 therapy in the past year (blow). *p<0.05. For patients with viral sequencing at multiple time points the last time point was utilized.

Relapsed or refractory lymphoma and recent systemic therapy were independent predictors of hospital re-admission for symptomatic COVID-19, the strongest individual treatments of which were anti-CD20 therapy (hazard ratio [HR] 6.57, 95% confidence interval [CI] 2.37–18.26) and bendamustine (HR 4.72, 95% CI 1.37–16.23); notably, BTK inhibitors were not associated with readmission (**Supplementary Figure 3b-c**). On multivariate analysis, only anti-CD20 therapy was independently associated with hospital readmission (HR 4.23, 95% CI 1.38–13.02) (**Figure 3a-b**). Among the laboratory parameters, a decrease in CD4^+^, but not CD8^+^ T-cells corelated with risk of re-admission (**Figure 3c**). Notably, there was no significant difference in initial inflammatory markers between patients who were or were not re-hospitalized for COVID-19 (**Supplementary Figure 3d**). Taken together, this data indicates that immunosuppression, and in particular loss of B- and CD4 T-cell immunity driven at least in part by B-cell directed therapies, is a risk factor for persistent symptomatic COVID-19 infection and suggests that CD4^+^ T-cell and/or B-cell function may contribute to complete viral eradication. Consistent with this hypothesis, patients who were ultimately re-hospitalized for COVID-19 exhibited an increased proportion of activated CD4^+^ and CD8^+^ T-cells, suggesting ongoing active infection in these patients (**Figure 3C**).

### Viral evolution in patients with persistent SARS-CoV-2 infection

Viral evolution, reinfection, and immune escape have been described in immunocompromised patients with COVID-19 infection.^8^ To determine whether there was any evidence of viral evolution, co-infection with different strains, or reinfection in lymphoma patients with persistent RNA detection and clinical symptoms, we analyzed 68 longitudinal samples from 18 of 19 persistently infected patients. Overall, there was >90% coverage for 78 analyzed samples. The timespan of the analyzed samples relative to the time of diagnosis is depicted in **Figure 4a**. The frequency of accumulation of the variants compared to the reference genome (NC_045512) and sustained vs. sporadic minor variant detection is demonstrated in **Figure 4a and Supplementary Figure 5**. Overall, single nucleotide variant analysis of consensus sequences (color-coded for the same patient) showed evidence of intra-host evolution suggesting ongoing infection. Conversely, there was no indication of reinfection or co-infection in any of the analyzed samples.

Finally, we computed the viral entropy, as defined in the Methods, across variant allele frequencies of all mutations which differed from the reference sequence, with a higher entropy indicating greater heterogeneity in the viral population. We observed higher viral entropy in patients with impaired CD8^+^ T-cell counts, but not in patients who had received anti-CD20 therapy in the past year (**Figure 4b**), suggesting that viral diversity may be constrained by an active CD8^+^ T-cell immune population and patients who harbor higher viral diversity during chronic infection have weaker adaptive responses.

## DISCUSSION

The unique contribution of this report is the description of persistent symptomatic COVID-19 in patients with lymphoid malignancy, a condition predisposed by previous anti-CD20 therapy and observed in 21% of anti-CD20 treated patients in our cohort. The syndrome manifests as a chronic protracted illness marked by slowly progressive or relapsing respiratory illness with progressive lower airway changes on CT imaging and persistent viral RNA detection. This complication affected 19/76 hospitalized patients with initial improvement and resulted in delay of cancer-directed therapy for 13/19 patients; 4/19 eventually died from the direct long-term sequelae of chronic COVID-19 infection. At a virologic level, we did not find any evidence of reinfection or co-infection with reactivation. Intra-host evolution of the virus suggested ongoing replication. Our analysis points to immune dysregulation characterized by combined loss of B-cell function and impaired CD4^+^ T-cell counts (CD4 <100 cells/µL) and compensatory innate immune and CD8^+^ T-cell activation as the basis for this chronic syndrome.

Multiple studies thus far have established a high mortality rate in cancer patients with acute COVID-19 infection^4,6,10,11^; the mechanistic explanations for this increased mortality rate have not been fully elucidated but may involve immune dysregulation and decreased functional status. The toxicity from COVID-19 infection clearly extends beyond the period of acute viral infection; the so-called “long COVID” syndrome is still an area of active exploration but is thought to be largely due to sequelae from prior infection rather than ongoing active infection.^12,13^ We had previously reported the presence of persistent PCR positivity with associated culturable virus in HSCT recipients lasting up to 3 months following initial COVID-19 infection.^14–18^

We now describe chronic persistent COVID-19 infection as a separate clinical entity in a significant proportion of hematologic malignancy patients and identify unique clinical and virologic characteristics associated with this syndrome. The immunologic factors associated with death during acute severe COVID-19 infection among heme cancer patients include increased acute phase reactants, decreased CD4^+^ and CD8^+^ lymphopenia and limited diversity in T-cell responses. These deficits are distinct from those observed in chronic SARS-CoV-2 infection, which is characterized by CD4^+^ lymphopenia and B-cell aplasia. Furthermore, the immune profile risk is also different from prolonged infection in immunocompromised patients with other severe respiratory virus illnesses such as influenza and respiratory syncytial virus where the extent of cell mediated immunosuppression drives viral persistence and is almost exclusively observed after HSCT.^19,20^ Overall, our findings suggest that distinct arms of the immune response play dominant roles in controlling acute infection due to a novel virus and influence permanent SARS-CoV-2 viral clearance.^21–24^ B-cell depletion due to anti-CD20 antibody therapy has been associated with viral persistence and reactivation especially in chronically infected hepatitis B patients following chemoimmunotherapy and an elevated risk of progressive multifocal leukoencephalopathy, herpes simplex virus and varicella-zoster virus has also been reported, but an association with prolonged viral shedding from respiratory viruses had yet to be described.^25,26^ The role of CD4^+^ T-cell depletion in viral persistence is less clear. CD4^+^ T-cell help is critical for both B-cell maturation and optimal CD8^+^ T-cell survival and function^27^; conversely B-cell depletion may impair naïve CD4^+^ T-cell homeostasis.^28^ Interestingly, greater intra-host viral diversity was observed mainly in patients with impaired CD8^+^ T-cell immunity. This may reflect greater evolution in the setting of partial immunity, as has been reported previously.^16^ Whether B-cell depletion alters the COVID-19-reactive CD8^+^ T-cell repertoire is an area of ongoing investigation.

There are several limitations in our present study. Laboratory markers, including flow cytometry data and viral sampling, were ordered at the discretion of the treating physician and were therefore not uniformly available across patients. Additionally, viral viability was not assessed in patients with persistent COVID-19 PCR positivity. Finally, the management of these patients was heterogeneous and physician dependent with rapidly emerging evidence. Despite this, the high volume of patients with hematologic malignancy and immune-depleting therapies at our institution enabled the identification of this novel syndrome.

In summary, the results of this work have significant implications for devising effective preventive and treatment strategies for COVID-infected patients with lymphoid malignancies. The high risk for adverse outcomes with acute COVID-19 in patients with lymphoid cancers is well recognized in multiple previous reports.^7,29^ We corroborate this finding among our large study cohort. Most importantly, we demonstrate the substantial morbidity and mortality associated with the long-term effects of chronic COVID-19 infection in lymphoma patients unable to attain adequate viral clearance and potentially posing a transmission risk to others. Future studies should elucidate the immunologic defects associated with this clinical phenomenon beyond the recent anti-CD20 treatment risk identified in our report. When taken in conjunction with emerging evidence on suboptimal COVID-19 vaccine response in anti-CD20 treated patients,^30–33^ our findings urge for prioritization of passive immunoprophylactic approaches in high-risk patients with lymphoid malignancy.

## Data Availability

Sharing of de-identified clinical data, upon request, will be considered by MSKCC's institutional review board (IRB).

## METHODS

### Study Setting and Methods

Memorial Sloan Kettering Cancer Center (MSKCC) is 514-bed tertiary care cancer with 22,417 annual admissions and 160,298 inpatient days in 2020. In 2019, MSKCC providers saw 18,800 patients with a new cancer diagnosis, including 946 patients with lymphoma and 335 and 235 cases of leukemia and myeloma, respectively. From 10 March 2020 until 28 February 2021, the current study included all consecutive adults with laboratory-confirmed SARS-CoV-2 infection and underlying hematologic malignancy. Identification of case-patients and their medical background and clinical course during COVID-19 illness were abstracted from electronic medical records. Longitudinal respiratory samples from 17 patients with lymphoid malignancies were retrieved for whole genomic sequencing.

### Laboratory Methods

#### SARS-CoV-2 RNA test

SARS-CoV-2 RNA was detected in nasopharyngeal swabs (NPS) or saliva samples using a laboratory-developed test as previously described.^4,34^ Testing was also performed using several commercial assays including the TaqPath™ COVID-19 Combo Kit (Thermo Fisher Scientific, Waltham, MA) targeting the N, S and ORF genes, the cobas® SARS-CoV-2 test (Roche Molecular Diagnostics, Indianapolis, IN) targeting the ORF1 a/b and E gene and the Xpert Xpress SARS-CoV-2 test (Cepheid, Sunnyvale, California) targeting the N and E genes. Samples were reported as positive per manufacturers’ instructions. The cycle threshold (Ct) value, a semi-quantitative estimate of the viral SARS-CoV-2 RNA load, was retrieved for all gene targets from each instrument record.

#### SARS-CoV-2 Whole Genome Sequencing (WGS)

WGS was performed on all samples with high enough viral load (i.e. Ct value < 30). Total viral nucleic acids were isolated from 200 µl of NPS or saliva samples on the KingFisher Flex Magnetic Particle Processor using the MagMAX™ Viral/Pathogen Nucleic Acid Isolation Kit (Thermo Fisher Scientific, Waltham, MA). Amplicon sequencing was performed following the Artic protocol with version 3 primers (Integrated DNA Technologies (IDT), Coralville, IA). Following cDNA synthesis and multiplexed PCR, libraries were prepared for the two amplicons pools using the Nextera XT DNA kit followed by sequencing on an Illumina Miseq platform (Illumina, San Diego, CA USA) as paired-end (2□×□150 base pair reads). We mapped the reads using STAR (version 2.7.7a) to the combined human (hg38 p.13, gencode release 36) and reference SARS-CoV-2 (NC_045512) genome^35^. Duplicates were removed using picard (version 2.24.2).^36^ Reads were subset to those mapping to the viral genome and subsequently post-processed as follows: first, ends of the reads were trimmed until the 10 bases closest to the end mapped to the reference with no mismatches. After that, the trimmed reads with overall identity to the reference less than 95% and reads shorter than 40 base pairs were discarded. In addition to that, if there were two mismatches to the reference within 5 bases of each other, the quality score for the corresponding bases was set to 2. After that the reads were written to FASTQ format and mapped to the SARS-CoV-2 genome using bwa (version 0.7.17-r1188).^37,38^ Single nucleotide variants (SNVs) were called using exactSNP program from the Subread package (v 2.0.0).^39^ When we encountered major variants with respect to the reference genome, we applied those variants and reran SNV calling with the updated reference. Settings which we modified from the default for SNV calling are -Q 7 -n 2 -s 20 -T 5. We increased the default p-value cutoff (changed the -Q parameter from the default value of 12 to 7) to account for the fact that the size of the SARS-CoV-2 genome is 10^5^ times smaller than the size of the human genome defaults are targeted at. When calling minor variants, we required the depth of coverage to be at least 50.

Consensus sequences of the viral genomes were assembled using bcftools.^40,41^ SARS-CoV-2 lineages were assigned using pangolin https://pangolin.cog-uk.io/.^42^

The Shannon entropy was computed for each variant site with the major and minor variant allele fractions:

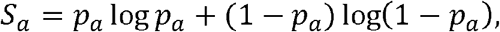

where *p*_*a*_ is the frequency of the minor allele at the site *a*. The total “viral entropy” per sequencing sample is the sum across the sites within that sample. This ignores the possible correlation between the polymorphic sites (i.e., minor variants observed at sites A and B can be present in the same clone or in two different clones), which generally cannot be determined using the short reads. On a per-patient basis, we utilized the sample viral entropy corresponding to the latest time point. Categorical tests were conducted using SciPy’s ttest_ind function.^43^ Linear and rank correlations to flow cytometry data were determined using SciPy’s pearsonr and spearmanr functions.^43^

#### Peripheral blood immunophenotyping

Peripheral blood was collected into BD Horizon Dri tubes (BD, Cat#625642). Immunophenotyping of peripheral blood mononuclear cells via flow cytometry was performed in the MSKCC clinical laboratory. The lymphocyte panel included CD45 FITC (BD, 340664, clone 2D1), CD56+16 PE (BD 340705, clone B73.1; BD 340724, clone NCAM 16.2), CD4 PerCP-Cy5.5 (BD 341653, clone SK3), CD45RA PC7 (BD 649457, clone L48), CD19 APC (BD 340722, clone SJ25C1), CD8 APC-H7 (BD 641409, clone SK1), and CD3 BV 421 (BD 562426, clone UCHT1). The naive/effector T panel included CD45 FITC (BD 340664, clone 2D1), CCR7 PE (BD 560765, clone 150503), CD4 PerCP-Cy5.5 (BD 341653, clone SK3), CD38 APC (BioLegend, 303510, clone HIT2), HLA-DR V500 (BD 561224, clone G46-6), CD45RA PC7 (BD 649457, clone L48), CD8 APC-H7 (BD 641409, clone SK1), and CD3 BV 421 (BD 562426, clone UCHT1). The immune phenotypes were based on NIH vaccine consensus panels and the Human Immunology Project. Samples were acquired on a BD Facs Canto using FACSDiva software. 8-color peripheral blood cytometry for annotation of T, B, and natural killer (NK) cell subsets was performed in 54.7% of hospitalized patients and 12-color peripheral blood flow cytometry for T-cell subset analysis was performed in 38.4%.

#### Data Availability

All sequences were uploaded to GISAID (accession numbers pending). The Memorial Sloan Kettering Cancer Center Institutional Review Board granted a Health Insurance Portability and Accountability Act waiver of authorization to conduct this study. Clinical data were abstracted from the electronic medical record into standardized case report forms. Clinical laboratory data were abstracted from the date closest to research blood collection. In addition to standard laboratory analyses performed on patients during acute COVID-19 infection, inflammatory markers including C-reactive protein, interleukin-6, D-dimer, ferritin, fibrinogen, and lactate dehydrogenase were measured in most inpatients. In addition, 8-color peripheral blood cytometry for annotation of T, B, and NK cell subsets was performed in 54.7% of hospitalized patients and 12-color peripheral blood flow cytometry for T-cell subset analysis was performed in 38.4%.

#### Statistical analysis

Descriptive statistics were used to summarize the data. Continuous variables were presented as means and standard deviations or as medians and ranges, as appropriate, and categorical variables were presented as counts and percentages. Overall survival (OS) was measured from the date of laboratory-confirmed COVID-19 to last follow-up or death. Data were censored on April 3, 2021. The probability of OS was estimated using the Kaplan–Meier method and differences compared using the log-rank test. Cox proportional hazard regression models were used to identify predictors of OS. Multivariate analysis using Cox proportional hazards model was used to identify the potential independent effects of those factors that attained a p-value of 0.1 in the univariate analysis. Risk factors for persistent SARS-CoV-2 positivity were analyzed using multivariate logistic regression; the final model was constructed based on a univariate p-value of 0.2. Differences in distributions of characteristics of patients were analyzed with the use of Pearson’s chi-square test for nominal or categorical variables and the Mann–Whitney test for ordinal or continuous variables. Analyses were performed with the use of SPSS software, version 26 and R version 4.0 (R Development Core Team, Vienna, Austria). Exact methods were used to calculate confidence intervals for the rate ratios. P values were calculated with the use of two-sided exact tests; a P value of less than 0.05 was considered to indicate statistical significance.

To determine whether there were distinct immunological responses to COVID-19 in our patient population, we identified 36 patients in whom at least 10% of laboratory analyses were performed and imputed missing data in these patients with the median of each feature (**Figure 2b**). We projected the multivariate laboratory data by conducting a Uniform Manifold Approximation and Project (UMAP) in two dimensions (**Supplementary Figure 2a**).^44^ Next, we computed the Earth Mover’s Distance (EMD) of each point in UMAP space to each other point.^45^ We clustered these distances and identified three clusters based on immunologic laboratory markers alone.

