## Supplementary Data for "Prolonged SARS-CoV-2 infection in patients with lymphoid malignancies"

**Supplementary Table 1. Demographic and clinical characteristics of 368 SARS-CoV-2 infected patients with hematologic malignancy and survival beyond 30 days from infection diagnosis.**

| **Characteristics** | **Total**  (N= 368) | **Percentage** |
| --- | --- | --- |
| **Age ≥65 years** | 118 | 32.1% |
| **Male sex** | 207 | 56.2% |
| **Race** |  |  |
| White | 245 | 66.6% |
| Black or African American | 56 | 15.2% |
| Other | 67 | 18.2% |
| **Ethnicity** |  |  |
| Hispanic or Latino | 72 | 19.6% |
| **Underlying Cancer** |  |  |
| Lymphoid~ | 200 | 54.3% |
| Myeloma | 85 | 23.1% |
| Leukemia | 68 | 18.5% |
| Myeloproliferative neoplasm | 15 | 4.1% |
| **Active treatment *** | 135 | 36.7% |
| **HSCT within 1 year** | 28 | 7.6% |
| **Anti-CD20 therapy within 1 year** | 67 | 18.2% |
| **Chronic lymphopenia^¶^** | 13 | 3.5% |
| **SARS-CoV-2 RNA positive ≥30 days** | 51 | 13.9% |

~ includes all lymphoma subtypes, Waldenström macroglobulinemia and chronic lymphocytic leukemia

* with systemic parenteral chemotherapy within last 6 months

¶ Absolute lymphocyte count <500 per microliter over five previous consecutive measurements prior to COVID-19 diagnosis

HSCT, hematopoietic stem cell transplantation

**Supplementary Table 2. Demographic and baseline clinical characteristics of lymphoma patients with COVID-19 infection.**

| Characteristic | n (%) or median (range) |
| --- | --- |
| Age, year | 61 (20–89) |
| Sex |  |
| Male | 119 (56%) |
| Female | 95 (44%) |
| Race / ethnicity |  |
| White | 136 (64%) |
| Hispanic or Latino | 34 (16%) |
| Black | 20 (9%) |
| Asian | 11 (5%) |
| Other / Unknown | 12 (6%) |
| Body mass index ≥30 kg/m^2^ | 85 (40%) |
| Cardiovascular comorbidity | 30 (14%) |
| Pulmonary comorbidity | 23 (11%) |
| Diabetes | 24 (11%) |
| Smoking status |  |
| Current | 9 (4%) |
| Former | 74 (35%) |
| Never | 127 (59%) |
| Unknown | 4 (2%) |
| Lymphoma subtype |  |
| Non-Hodgkin lymphoma | 183 (86%) |
| Mature B cell lymphoma | 165 (77%) |
| Mature T cell lymphoma | 18 (8%) |
| Hodgkin lymphoma | 31 (14%) |
| Disease status at time of COVID-19 diagnosis |  |
| Relapsed or refractory disease | 85 (40%) |
| Active treatment or management | 79 (37%) |
| Remission | 53 (25%) |
| Surveillance | 82 (38%) |
| Treatment history |  |
| No. prior therapies | 1 (0–11) |
| Any systemic therapy within 1 year | 107 (50%) |
| Anti-CD20 within 1 year | 69 (32%) |
| BTK inhibitor within 1 year | 21 (10%) |
| Bendamustine within 1 year | 10 (5%) |
| Autologous or allogeneic HSCT within 2 years | 15 (7%) |
| CD19-targeted CAR T-cell therapy within 1 year | 6 (3%) |

BTK, Bruton tyrosine kinase; CAR, chimeric antigen receptor; HSCT, hematopoietic stem cell transplantation

**Supplementary Table 3. Inpatient management of lymphoma patients with COVID-19 infection.**

| **Intervention** | **n (%)** |
| --- | --- |
| Supplemental oxygen | 73 (72%) |
| Mechanical ventilation | 22 (22%) |
| Systemic corticosteroids | 53 (52%) |
| Convalescent plasma | 45 (44%) |
| Hydroxychloroquine | 40 (39%) |
| Remdesivir | 38 (37%) |
| N-acetylcysteine | 17 (17%) |
| Intravenous immune globulin | 9 (9%) |
| Tocilizumab | 7 (7%) |
| Anakinra | 1 (1%) |

**Supplementary Table 4. Clinical outcomes of lymphoma patients with COVID-19 infection.**

| **Characteristic** | | ***n* (%) or median (range)** |
| --- | --- | --- |
| **Hospitalization** | | 102 (47%) |
|  | Duration of hospitalization (days) | 11 (1–132) |
|  | ICU admission | 32 (31%) |
|  | Hospital readmission | 19 (19%) |
|  | Remains hospitalized | 1 (1%) |
|  | Lost to follow-up | 1 (1%) |
| **Recovered/improved from COVID-19** | | 185 (86%) |
|  | Lymphoma therapy modification | 49 (26%) |
|  | Disease status |  |
|  | Remission or surveillance | 144 (78%) |
|  | Active treatment | 27 (15%) |
|  | Progression of disease | 14 (8%) |
| **Deaths** | | 33 (15%) |
|  | COVID-19 | 27 (82%) |
|  | Lymphoma progression | 3 (9%) |
|  | Other infection | 2 (6%) |
|  | Secondary malignancy | 1 (3%) |

**Supplementary Figure 1. Predictors of mortality in lymphoma patients with acute COVID-19 infection.** a) Univariate analysis of clinical risk factors predicting mortality attributable to COVID-19 infection in patients with lymphoma. Statistically significant risk factors highlighted in red. b) Laboratory markers in patients who were alive or dead following acute COVID-19 infection. *p<0.05.

**Supplementary Figure 2. Unbiased clustering reveals distinct immunological patterns in lymphoma patients with COVID-19 infection.** a) UMAP projection of 36 patients with lymphoma in whom at least 10% of all laboratory analyses were conducted. Clusters identified based on Earth Mover’s Distance (EMD) of each patient relative to each other in UMAP space is shown. b) Differences in immunologic parameters between three EMD clusters shown in a). *p<0.05. **p<0.01. ***p<0.001.

**Supplementary Figure 3. Predictors of COVID-19-specific re-hospitalization in lymphoma patients with prior COVID-19 infection.** a) Inclusion criteria for cohorts used for analysis in Figure 3 and Supplementary Figure 3. b) Univariate analysis of clinical risk factors predicting re-hospitalization attributable to COVID-19 infection in patients with lymphoma and prior hospitalization attributable to COVID-19. Statistically significant risk factors highlighted in red. c) Kaplan-Meier curve displaying probability of re-hospitalization for COVID-19 in patients with lymphoma based on whether patients were undergoing active lymphoma-directed therapy at the time of first known documented SARS-CoV-2 PCR positivity. d) Laboratory markers in patients with lymphoma who either died from acute COVID-19 infection, survived acute infection and were not re-admitted to the hospital, or survived acute infection and were re-admitted to the hospital for COVID-19 infection. **p<0.01. ***p<0.001. ****p<00001.

**Supplementary Figure 4. Radiographic findings in hematologic malignancy patients with persistent SARS-CoV-2 infection.** Serial CT images for Patients E, H, M (from Table 2) over three separate time points from initial diagnosis. Patient E with waxing and waning ground glass opacities and patchy infiltrates, later characterized by increasing consolidation. Patient H with waxing and waning ground glass opacities with eventual clearing. Patient M with waxing and waning ground glass opacities with coarsening over time.

**Supplementary Figure 5. Major and minor SARS-CoV-2 genome variants in longitudinal samples from patients with persistent COVID-19 infection.** Major variants in SARS-CoV-2 sequences from 17 patients with longitudinal data. Samples from the same patient are grouped together and highlighted with the same background color. White gaps indicate regions with low sequencing coverage. Major variants are computed w.r.t. the reference sequence. Vertical lines indicate the location of the variants in the SARS-CoV-2 genome. The height of the line indicates the frequency of the variant in the viral population.
