## Supplementary Figures for "Prolonged SARS-CoV-2 infection in patients with lymphoid malignancies"

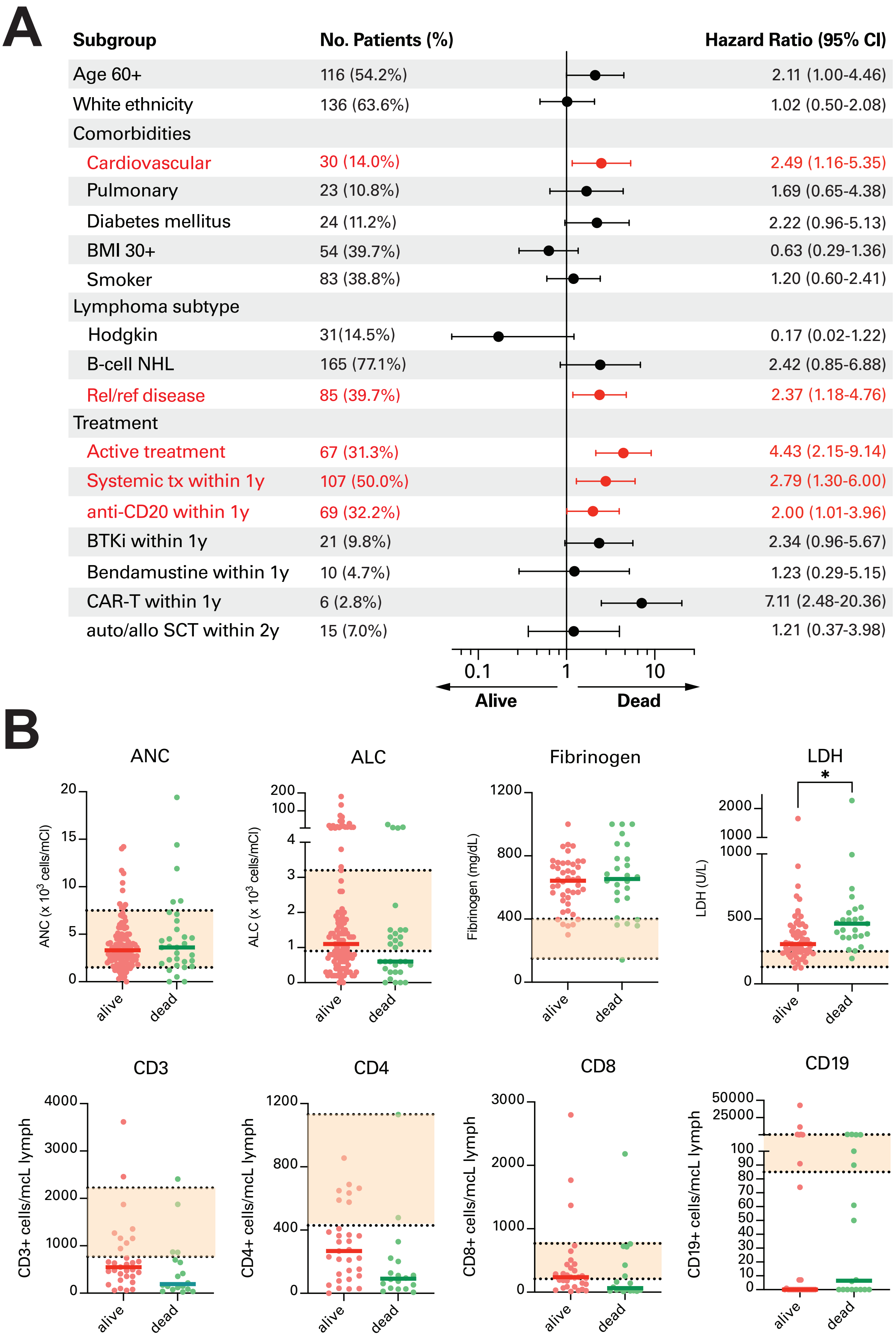


Supplementary Figure 1

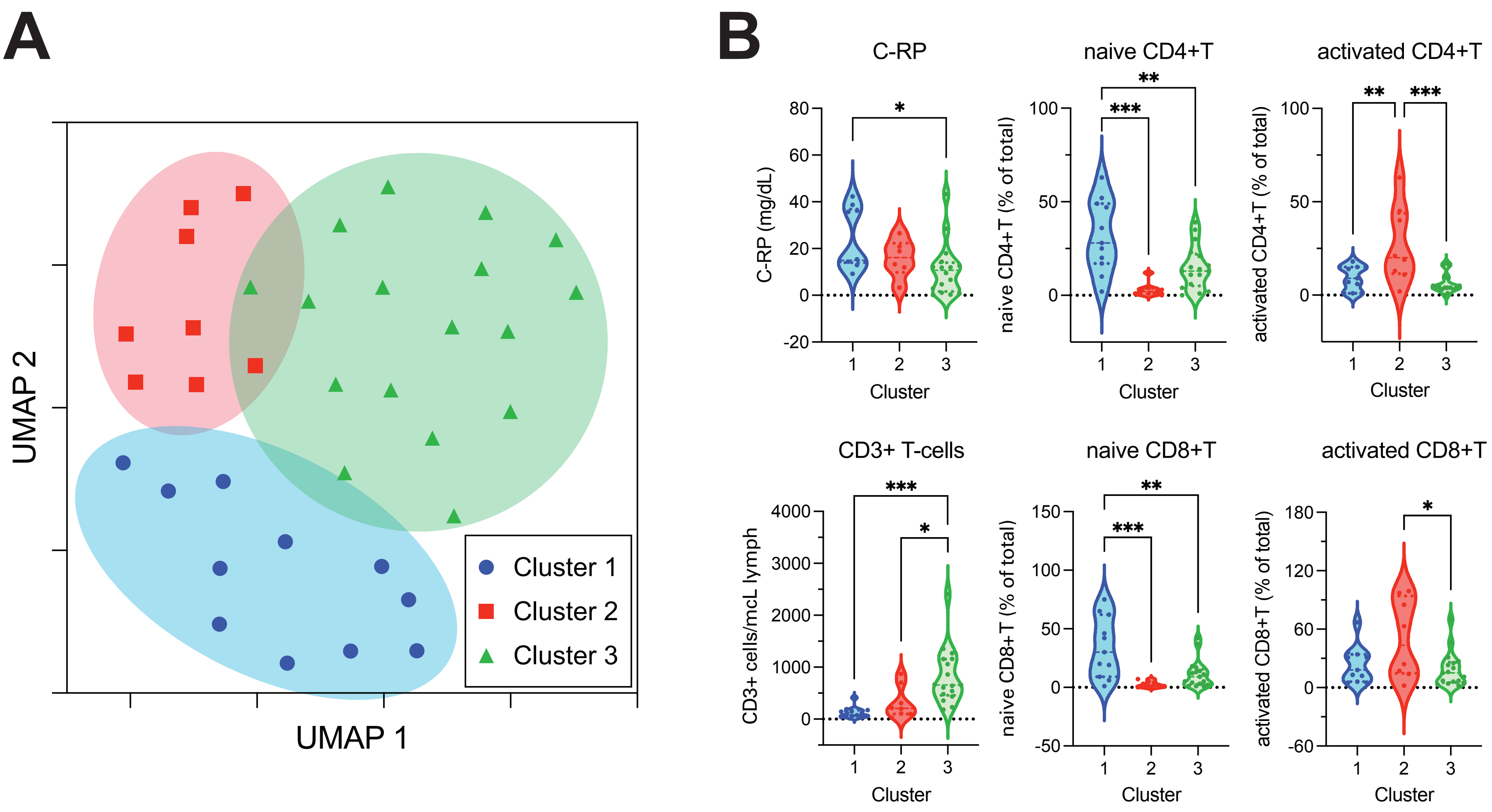


**Supplementary Figure 2**

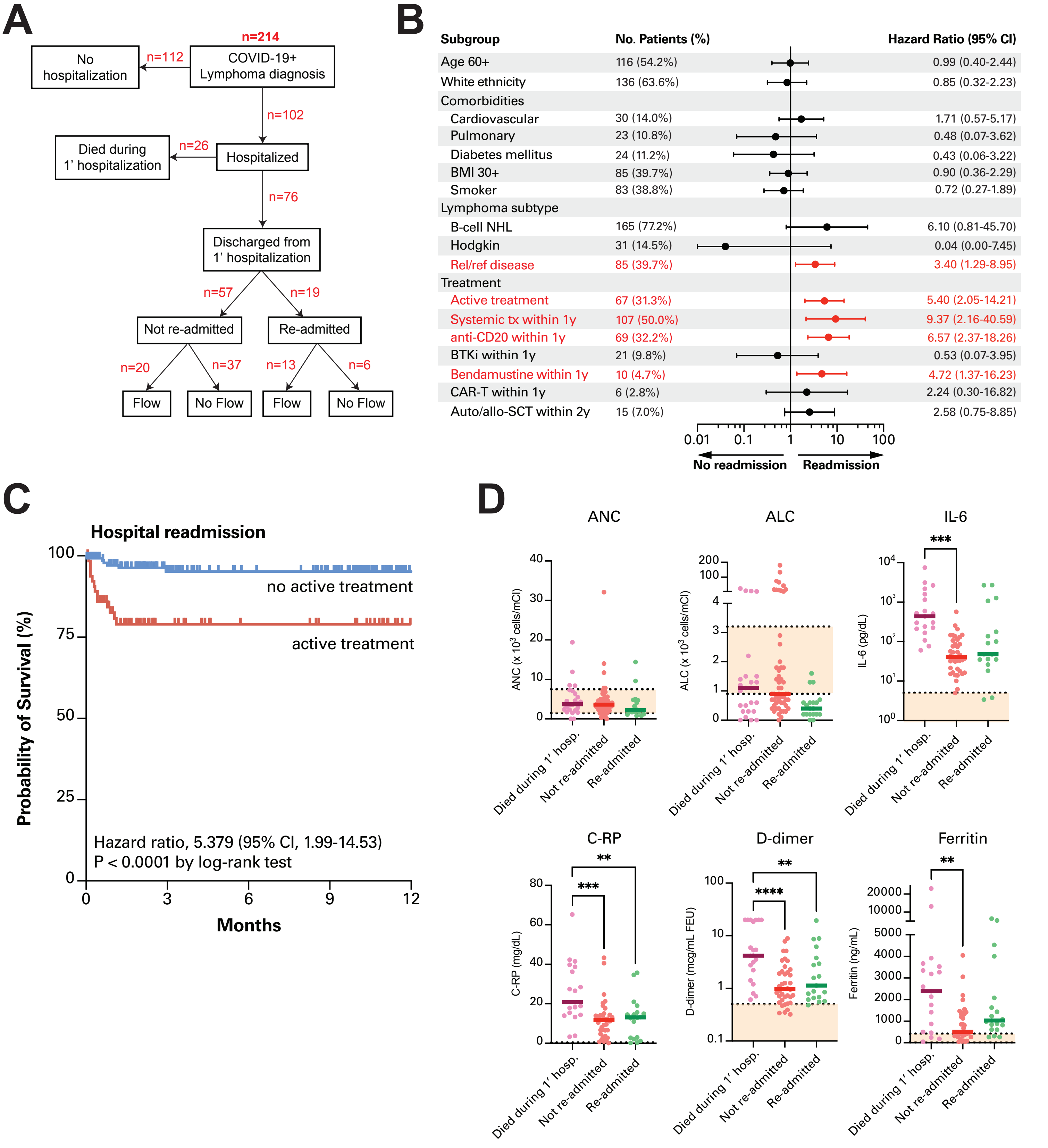


Supplementary Figure 3

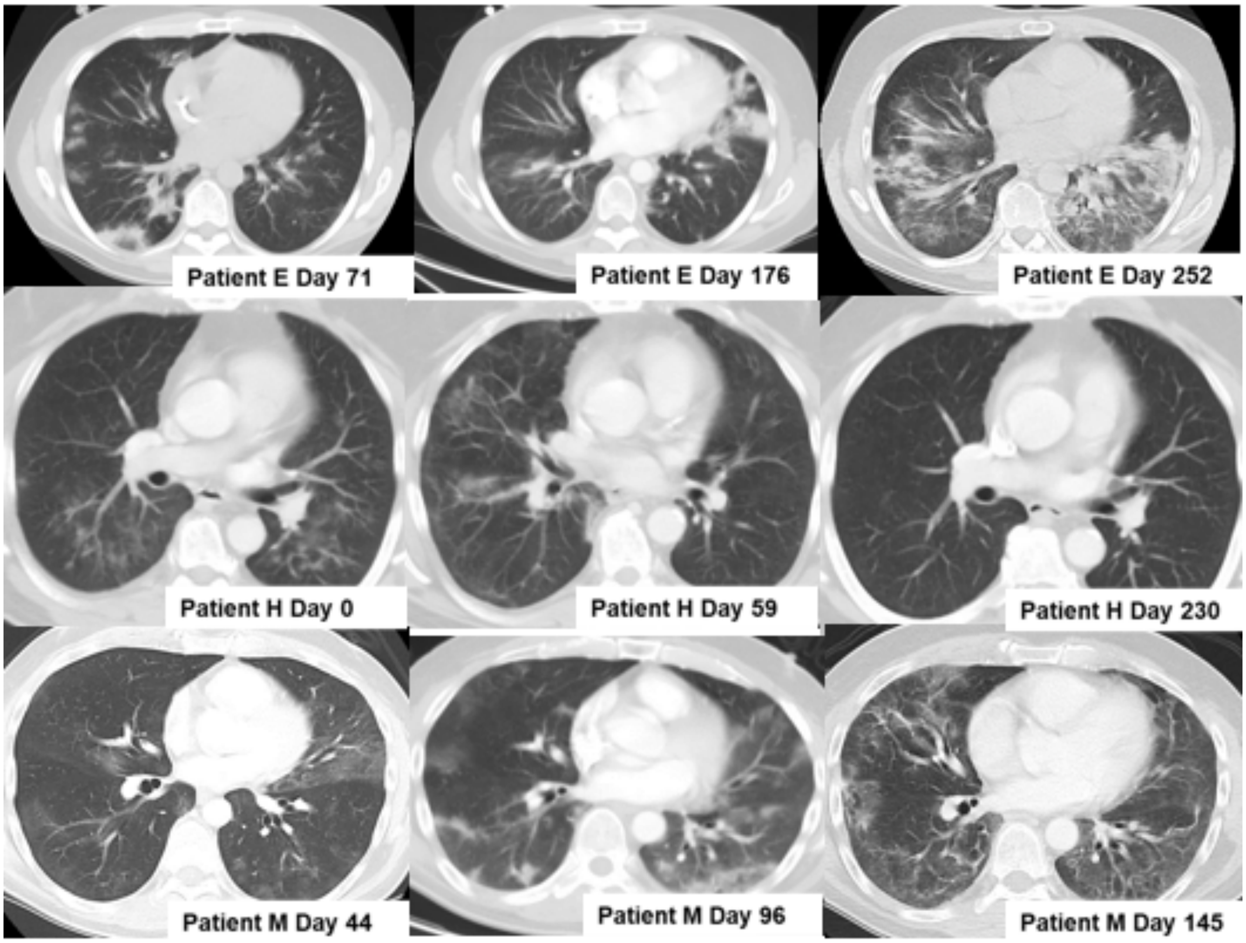


Supplementary Figure 4.

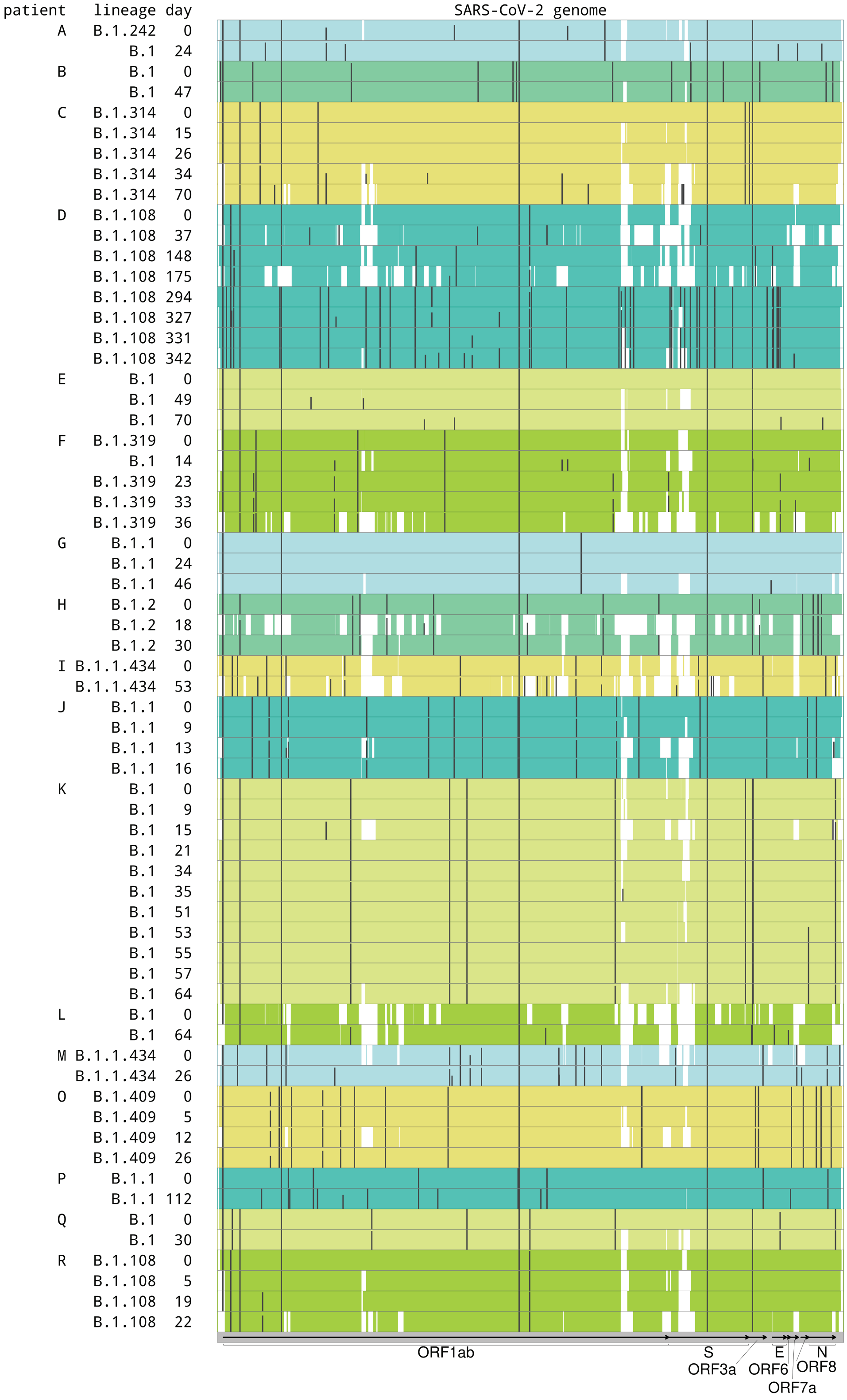


**Supplementary Figure 5**
